# Pump-Free Patient-Derived Human Proximal Tubule Microphysiological System for Modeling Flow-Dependent Epithelial Maturation and Cisplatin Injury

**DOI:** 10.64898/2026.06.18.26355848

**Authors:** Yuta Sekiguchi, Ayumi Suzuki, Yuki Nakao, Takeshi Hori, Makiko Mori, Afreen Fatima Mirza, Ryota Shindoh, Iori Morita, Shintaro Mandai, Tamami Fujiki, Hiroaki Kikuchi, Yohei Arai, Fumiaki Ando, Koichiro Susa, Takayasu Mori, Yuma Waseda, Soichiro Yoshida, Yasuhisa Fujii, Eisei Sohara, Yuji Nashimoto, Hirokazu Kaji, Yutaro Mori

## Abstract

Recent initiatives by the U.S. Food and Drug Administration and the National Institutes of Health to reduce animal testing in drug development have highlighted the need for *in vitro* platforms that better recapitulate human biology for preclinical safety assessment. Drug-induced nephrotoxicity remains a major cause of drug attrition, underscoring the need for human-relevant kidney models. To address this, a pump-free human patient-derived proximal tubule microphysiological system was developed by integrating human renal proximal tubular epithelial cells (hRPTECs), isolated from non-tumorous nephrectomy cortex, with a porous membrane-based microfluidic device. Expanded hRPTECs were cultured for 10 days under static conditions or rocker-driven shear stress approximating physiological proximal tubular flow. Shear stress increased epithelial density, enhanced proximal tubule marker expression (Na^+^/K^+^-ATPase and aquaporin-1), and improved Zonula occludens-1 and occludin localization. Bulk RNA sequencing demonstrated transcriptomic changes associated with enhanced apical maturation and epithelial signature. In cisplatin-induced injury assays, shear-conditioned epithelia exhibited reduced cell density and increased γH2AX staining, indicating greater sensitivity to nephrotoxicity. These findings demonstrate that rocker-driven shear stress promotes epithelial maturation in patient-derived hRPTECs. The pump-free human patient-derived proximal tubule microphysiological system offers a practical, scalable, and physiologically relevant platform for modeling flow-dependent proximal tubule biology and assessing human-relevant nephrotoxicity.

## 1. Introduction

In recent years, there has been a global movement toward reducing the use of experimental animals in drug development and biomedical research [1]. This shift has been reflected in evolving regulatory and policy frameworks, including the enactment of the U.S. Food and Drug Administration Modernization Act 3.0 [2], which permits alternatives to traditional preclinical animal testing prior to clinical trials. Although these initiatives are partly motivated by ethical considerations related to animal welfare, they are also driven by longstanding scientific concerns. Substantial interspecies differences between humans and commonly used laboratory animals limit the translational relevance of preclinical findings, as results observed in animal models often fail to predict human outcomes [3, 4]. Consequently, high attrition rates in drug development remain a major challenge, with many candidates that demonstrate promising efficacy and safety in preclinical studies ultimately failing during clinical trials. Indeed, multiple analyses have estimated that fewer than 10% of drug candidates entering clinical development ultimately receive regulatory approval [5–8].

In response, substantial efforts have been devoted to the development of alternative nonclinical approaches, including predictive modeling**-**based artificial intelligence and human-derived *in vitro* platforms, such as microphysiological systems (MPSs). Among various systems, the kidney is both a critical target for drug toxicity and one of the most challenging organs to faithfully recapitulate *in vitro*. Drug-induced nephrotoxicity is a major contributor to drug attrition during development and frequently results in the discontinuation of otherwise promising compounds [9]. These challenges underscore the urgent need for robust and human-relevant *in vitro* platforms capable of evaluating renal toxicity and pharmacological efficacy.

To address this need, several strategies have been developed, including induced pluripotent stem cell-derived kidney organoids [10] and MPSs [11]. Although induced pluripotent stem cell-derived organoids provide valuable models for investigating kidney development and disease, they require lengthy differentiation protocols and remain technically complex. In addition, many previously reported kidney MPS platforms rely on immortalized cell lines or non-patient-derived cell sources, which may not fully recapitulate the structural and functional characteristics of native human kidney tissue [12]. Primary human proximal tubular epithelial cells (PTECs) represent a promising alternative for the development of more physiologically relevant kidney models. However, the extent to which patient-derived PTECs undergo epithelial maturation and maintain phenotypic stability under sustained pump-free microfluidic shear stress has yet to be fully elucidated.

Here, we developed a pump-free, patient-derived proximal tubule MPS (hPT-MPS) using human renal PTECs (hRPTECs) isolated from surgically resected kidney tissue. By applying rocker-driven microfluidic shear stress, we investigated its impact on epithelial maturation and the response to cisplatin-induced tubular injury. This platform provides a practical and scalable framework for modeling flow-dependent proximal tubule biology in a human-derived setting.

## 2. Methods

### 2.1. Establishment of Human Renal Proximal Tubule Epithelial Cells

Human kidney specimens were collected from patients undergoing clinically indicated nephrectomy at the Institute of Science Tokyo Hospital, Tokyo, Japan. The study protocol was approved by the Institutional Review Board of the Institute of Science Tokyo Hospital (approval no. M2022-005). hRPTECs were isolated from the non-tumorous cortical regions of kidneys resected for renal cell carcinoma or urothelial malignancies using a modified version of a previously established method [13]. Briefly, renal cortical tissue was finely minced and enzymatically digested with collagenase type II (1.0 mg/mL) (Worthington Biochemical, NJ, USA). The enzymatic reaction was terminated by the addition of fetal bovine serum, and the digested tissue was resuspended in hRPTEC growth medium, consisting of Dulbecco’s Modified Eagle Medium/Nutrient Mixture F-12 (Nacalai Tesque, Kyoto, Japan), supplemented with bovine serum albumin (Nacalai Tesque), antibiotic–antimycotic solution (Thermo-Fisher Scientific, MA, USA), hydrocortisone (Thermo-Fisher Scientific, MA, USA), insulin-transferrin-selenium liquid media supplement (Sigma-Aldrich, MO, USA), and human recombinant epidermal growth factor (Thermo-Fisher Scientific, MA, USA). Cells were cultured for 7–10 days at 37°C in a humidified atmosphere containing 5% CO_2_ in an incubator.

### 2.2. Fabrication of the Microfluidic Chip Mold

Microfluidic chip molds were designed using computer-aided design software (Autodesk Fusion 360, Autodesk Inc., USA) and fabricated using a resin-based three-dimensional printer (ELEGOO Saturn 4 Ultra 16K, ELEGOO, China). Two types of straight microchannels were designed: with dimensions of 1 mm (width) × 15 mm (length) × 1 mm (height) and 1 mm (width) × 15 mm (length) × 100 µm (height), with different mold heights used to generate distinct upper and lower channel depths. The printable resin was prepared by supplementing polyethylene glycol diacrylate (Sigma-Aldrich, USA) with 1% (w/w) Omirad 819 (Toyotsu Chemiplas Corporation, Japan) and 1% (w/w) 2-isopropylthioxanthone (Tokyo Chemical Industry, Japan). The resin mixture was protected from light by wrapping the container with aluminum foil and stirred at 40°C and 600 rpm overnight to ensure complete dissolution of the photoinitiators. The prepared resin was then loaded into the printer reservoir and printed according to the manufacturer’s instructions. After printing, the molds were removed from the build platform and immersed in 99.5% ethanol (FUJIFILM Wako Pure Chemical Corporation, Japan) to remove residual uncured resin. The molds were subsequently post-cured by ultraviolet irradiation (FV-209B, FIFTY VISIONARY) for 2 minutes on each of the top and bottom surfaces. Finally, the molds were incubated in a 70°C oven overnight to ensure complete polymerization.

### 2.3. Microfluidic Device

Polydimethylsiloxane (PDMS) chips were fabricated using SILPOT™ 184 (Dow Toray Co., Ltd., Japan). The silicone elastomer base and curing agent were mixed at a 10:1 (w/w) ratio, degassed under vacuum for at least 15 min, cast onto the fabricated molds, and cured at 70°C for at least 2 h. Following curing, the PDMS layers were carefully peeled from the molds, and 4-mm inlet and outlet ports were created using biopsy punches. Semipermeable polyester (PET) membranes (0.4 µm pore size) were subsequently surface-functionalized by silanization. Briefly, the polyester membranes were treated with oxygen plasma and immersed in a solution of 3-aminopropyltriethoxysilane (1:19 v/v in purified water) at 80°C for 30 minutes, followed by washing and air-drying. The functionalized membranes were then trimmed to fit the channel dimensions and bonded to the oxygen plasma-treated upper PDMS layer. For device assembly, a thin PDMS adhesive layer (base curing agent = 10:3, w/w) was spin-coated onto a glass slide treated with trichloro (1H, 1H, 2H, 2H-perfluorooctyl) silane (Sigma-Aldrich, USA) and subsequently transferred to the lower PDMS channel surface. The upper and lower PDMS layers were carefully aligned and bonded, followed by overnight curing at approximately 45°C to produce a sealed multilayer microfluidic device.

### 2.4. Shear Stress Calculation

The physiological shear stress acting on proximal tubular epithelial cells was estimated theoretically based on reported human renal physiological parameters. The total glomerular filtration rate in healthy adults was assumed to be approximately 100 mL/min for both kidneys combined. Assuming that each kidney contains approximately 1.0 × 10^6^ nephrons per kidney (2.0 × 10^6^ total nephrons), the single-nephron volumetric flow rate (Q) at the entrance of the proximal tubule was calculated as:

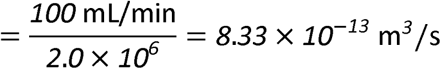

The proximal tubule was modeled as a cylindrical conduit with an average inner diameter of 25 µm (radius, *r* = 12.5 µm). For the proximal tubule, the shear stress was estimated using the following equation:

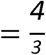

Where μ is the dynamic viscosity of primary urine (assumed to be 0.0008595 Pa·s). Based on the calculation, the shear stress in the proximal tubules under physiological conditions was estimated to be 4.67 dyn/cm^2^.

### 2.5. Shear Stress Setting

Devices were placed on a laboratory rocker to generate gravity-driven bidirectional perfusion. The maximum shear stress (τ) under rocker-induced flow was estimated using the following equation:

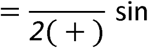

where *w* is the channel width (1.0 × 10^−3^ m), *h* is the channel height (1.0 × 10^−3^ m), ρ is the fluid density (1,000 kg/m^3^), *g* is the gravitational acceleration (9.8 m/s^2^), and θ is the tilting angle. When the device was tilted at 11°, the theoretical maximum shear stress was estimated to be 4.67 dyn/cm^2^. Based on the physiological estimation described in the shear stress calculation section, 4.67 dyn/cm² was considered to approximate *in vivo* proximal tubular shear stress in this study. Based on this calculation, we fabricated two device geometries to generate distinct shear stress conditions: a high-shear stress device with a channel height of 1 mm, corresponding to an estimated maximum shear stress of 4.67 dyn/cm², and a low-shear stress device with a channel height of 100 μm, corresponding to an estimated maximum shear stress of 0.85 dyn/cm².

### 2.6. Cell Culture

hRPTECs at several passages were seeded into the microfluidic device at a density of 1.0 × 10^6^ cells/mL in channels measuring 1 mm in width and 1 mm in height (high-shear condition). For devices with smaller channels measuring 1 mm in width and 100 μm in height (low-shear condition), cells were seeded at a 10-fold higher concentration to account for the 10-fold reduced channel volume. After 24 hours of static (no shear stress) culture, shear stress was applied using a rocker system, and the cells were subsequently cultured for 10 days in a humidified CO_2_ incubator.

### 2.7. Bulk RNA-Sequencing Analysis

Cells were cultured under high-shear stress (4.67 dyn/cm²) or static (no shear stress) conditions and harvested on Day 11. Total RNA was extracted using Sepasol (Nacalai Tesque, Japan) and further purified using the Direct-zol RNA Miniprep Plus Kit (Zymo Research, CA, USA). Samples (*n* = 4 per group) with RNA integrity number > 7.5 were processed by Rhelixa Inc. using NEBNext library preparation kits and sequenced on the NovaSeq X Plus platform. Libraries were prepared in a non-strand-specific manner. Raw sequencing reads were quality-trimmed using fastp and quantified using Salmon (GENCODE v45 (GRCh38 reference genome)). Detailed sequencing parameters are provided in the Supplementary Methods. Differentially expressed genes were identified using DESeq2. Principal component analysis and heatmaps were generated in RStudio. Gene set enrichment analysis was subsequently conducted using the Hallmark gene sets from the molecular signatures database based on a ranked gene list derived from DESeq2 output. Raw count data were analyzed using DESeq2, followed by variance-stabilizing transformation (VST). Box plots comparing gene expression between conditions were generated using VST-transformed expression values. Signature scores were also calculated from VST-transformed expression values. The apical maturation score was computed using eight genes: *LRP2, CUBN, SLC5A2, AQP1, EZR, ANPEP, RAB11A*, and *SNX32*. *VIL1* was initially considered but excluded from the final scoring due to lack of variance across samples. The epithelial score was calculated using eight epithelial marker genes: *EPCAM, CDH1, KRT8, KRT18, CLDN2, CLDN10, OCLN*, and *TJP1*.

### 2.8. Immunofluorescence Staining

Cells within the microfluidic device were washed three times with phosphate-buffered saline (PBS) and fixed with 4% paraformaldehyde in PBS. After removal of PBS from the device, cells were permeabilized with 0.1% Triton X-100 in PBS for 10 minutes at room temperature, followed by blocking with 3% bovine serum albumin in PBS for 10 minutes at room temperature. After removal of the blocking solution, the channels were filled with primary antibody solution and incubated overnight at 4°C. Following primary antibody incubation, the channels were washed three times with PBS and incubated with fluorescently labeled secondary antibodies for 30 minutes at room temperature in the dark. After two additional PBS washes, the device was disassembled, and the PET membrane was carefully retrieved. The membrane was mounted onto a glass slide using mounting medium containing Hoechst (P36981, Thermo Fisher Scientific, MA, USA), covered with a coverslip, and sealed to prevent leakage. The following primary anti-aquaporin-1 (AQP1) antibody (1:100) (SC-32737, Santa Cruz Biotechnology), rabbit anti-zonula occludens-1 (ZO-1) antibody (1:200) (21773-1-AP, Proteintech), mouse anti-occludin antibody (1:200) (OC-3F10, Invitrogen), and rabbit anti-phospho-Histone H2AX (Ser139) antibody (1:200) (9718S, Cell Signaling Technology). Fluorescence images were acquired using standard and/or confocal fluorescence microscopy (Eclipse Ti2, Nikon, Japan). Image analysis was performed using NIS-Elements Advanced Research software and ImageJ Fiji (https://imagej.net/software/fiji/downloads). For quantification, three randomly selected fields per sample were analyzed. Due to substantial inter-batch variability in staining intensity, image acquisition and quantification parameters were optimized independently for each batch. To minimize batch effects, all measurements were normalized to the mean value of the corresponding control group within each batch.

### 2.9. Assessment of Cisplatin-Induced Nephrotoxic Response

Cells were cultured under high-shear or static (no shear stress) conditions until Day 11. On Day 11, cisplatin was added at final concentrations of 0, 12.5, 25, and 50 μM, followed by 48 hours of incubation. Cells were then fixed and subjected to immunofluorescence staining. For quantification, three randomly selected fields per sample were analyzed. Fluorescent-positive areas were quantified using ImageJ/Fiji software by measuring the total fluorescent area.

### 2.10. Cell Number Quantification

For quantification of cell number in **Figure 2C**, phase-contrast images were acquired at 400× magnification on Day 11. Cells were manually counted in randomly selected microscopic fields under static (no shear stress), low-shear, and high-shear conditions. Cell number per field was used for quantitative analysis, with nine fields analyzed per condition.

### 2.11. Quantification and Statistical Analysis

The number of samples analyzed in each experiment is provided in the corresponding figure legends. Statistical significance between multiple groups was assessed using one-way analysis of variance followed by Tukey’s multiple comparisons test. For two-group comparisons of RNA-seq data, the two-sided Wilcoxon rank-sum test was used. To assess the effects of two independent factors on cisplatin response, a two-way analysis of variance followed by Tukey’s multiple comparisons test was performed. All statistical analyses were conducted using GraphPad Prism (GraphPad Software Inc., San Diego, CA, USA) or RStudio (https://cran.r-project.org/bin/windows/base/). Statistical significance is reported as follows (ns, not significant; \**p* < 0.05; \*\**p* < 0.01; \*\*\**p* < 0.001; \*\**p* < 0.0001).

## 3. Results

### 3.1. Establishment of a Human Patient-Derived Proximal Tubule Microphysiological System

A patient-derived hPT-MPS was developed by integrating primary hRPTECs with a porous membrane-based microfluidic device and rocker-driven shear stress (**Figure 1A**).

**Figure 1.**
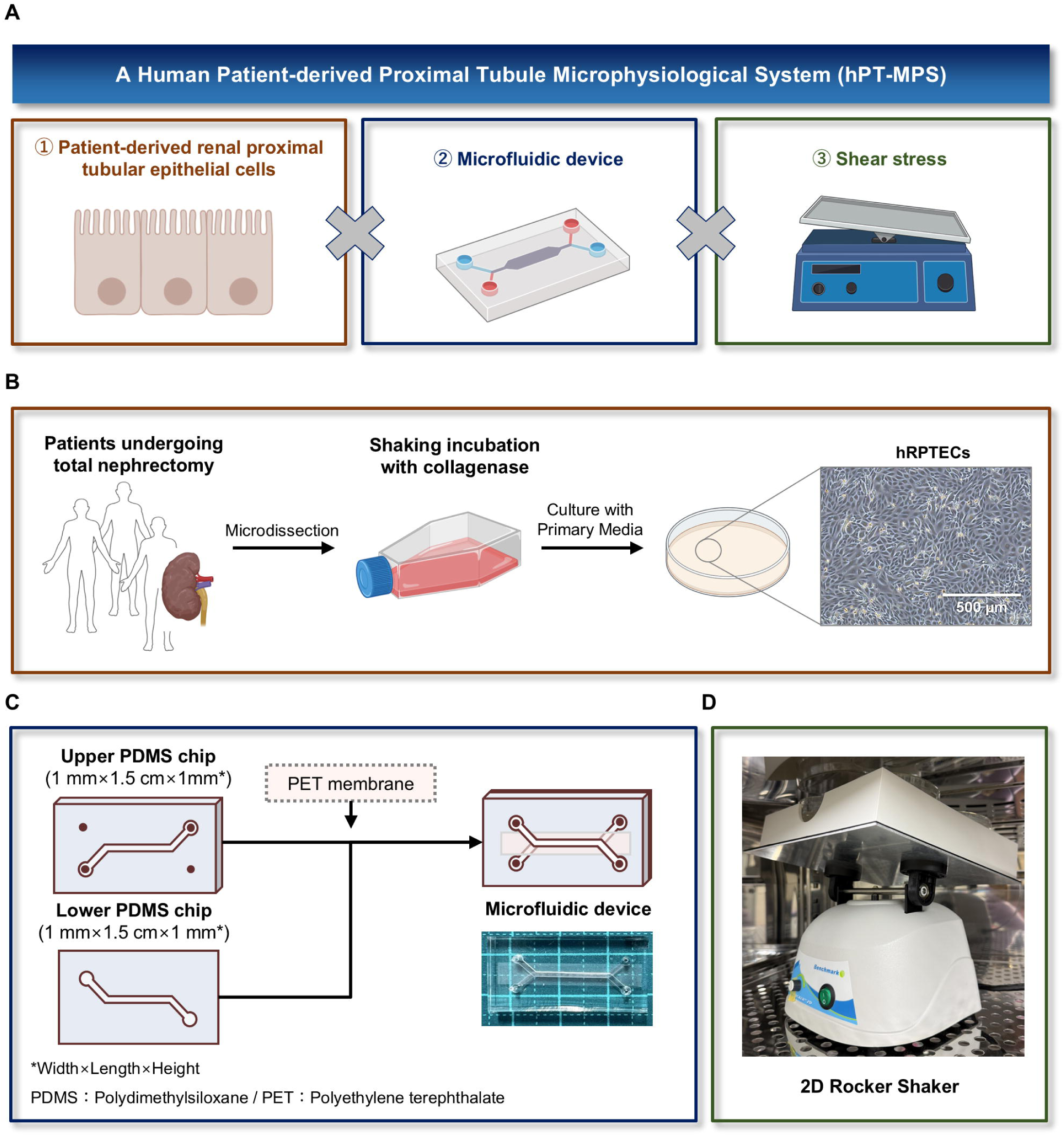
Establishment of a pump-free patient-derived human proximal tubule microphysiological system (hPT-MPS). **(A)** Schematic overview of hPT-MPS integrating patient-derived renal proximal tubular epithelial cells (hRPTECs), a microfluidic device, and shear stress. **(B)** Schematic workflow for the isolation and expansion of hRPTECs from nephrectomy specimens. Scale bar, 500 μm. **(C)** Schematic illustration of the microfluidic device comprising upper and lower polydimethylsiloxane (PDMS) layers separated by a polyethylene terephthalate (PET) membrane. **(D)** Photograph of the two-dimensional (2D) rocker platform used to generate pump-free bidirectional flow. Figure 1 was created in part using <u>BioRender.com</u>.

Primary hRPTECs were isolated from the non-tumorous cortical regions of nephrectomy specimens obtained from patients undergoing total nephrectomy. Following microdissection, tissues were enzymatically dissociated with collagenase and cultured in primary growth medium to promote epithelial outgrowth (**Figure 1B**). The resulting cells exhibited a characteristic cobblestone-like morphology consistent with an epithelial phenotype. To further assess proximal tubule-associated transcriptional features, the expression of previously reported 193-gene proximal tubule signatures was assessed [14]. On Day 11, hRPTECs cultured in the microfluidic device expressed approximately 60% of these proximal tubule-associated genes at transcripts per million values > 15 under both no-shear and shear stress conditions (**Supplementary Figure S1**). Although the analytical approaches were not identical, this proportion exceeded those reported for several conventional renal cell lines (approximately 15%–45%) and was comparable to or higher than that reported for primary mouse tubular epithelial cells (approximately 50%) [14]. These findings suggest that patient-derived hRPTECs retained a substantial proportion of proximal tubule-associated transcriptional features under the experimental culture conditions.

The microfluidic device comprised upper and lower PDMS layers separated by a porous PET membrane (**Figure 1C**). Continuous bidirectional flow was generated by placing the device on a two-dimensional rocker platform, enabling the pump-free application of shear stress within the microchannel (**Figure 1D**). Collectively, these components enabled the establishment of a patient-derived hPT-MPS that integrates primary hRPTECs, a porous membrane-based microfluidic device, and rocker-driven shear stress.

### 3.2. Physiological Shear Stress Increases Cell Number and Density within the Human Patient-Derived Proximal Tubule Microphysiological System

Next, the effects of shear stress on hRPTEC behavior within the microfluidic device were investigated. Following cell seeding, hRPTECs were cultured for up to 11 days under static (no shear stress), low-shear stress (0.85 dyn/cm²) or high-shear stress (4.67 dyn/cm²) conditions **(Figure 2A**).

**Figure 2.**
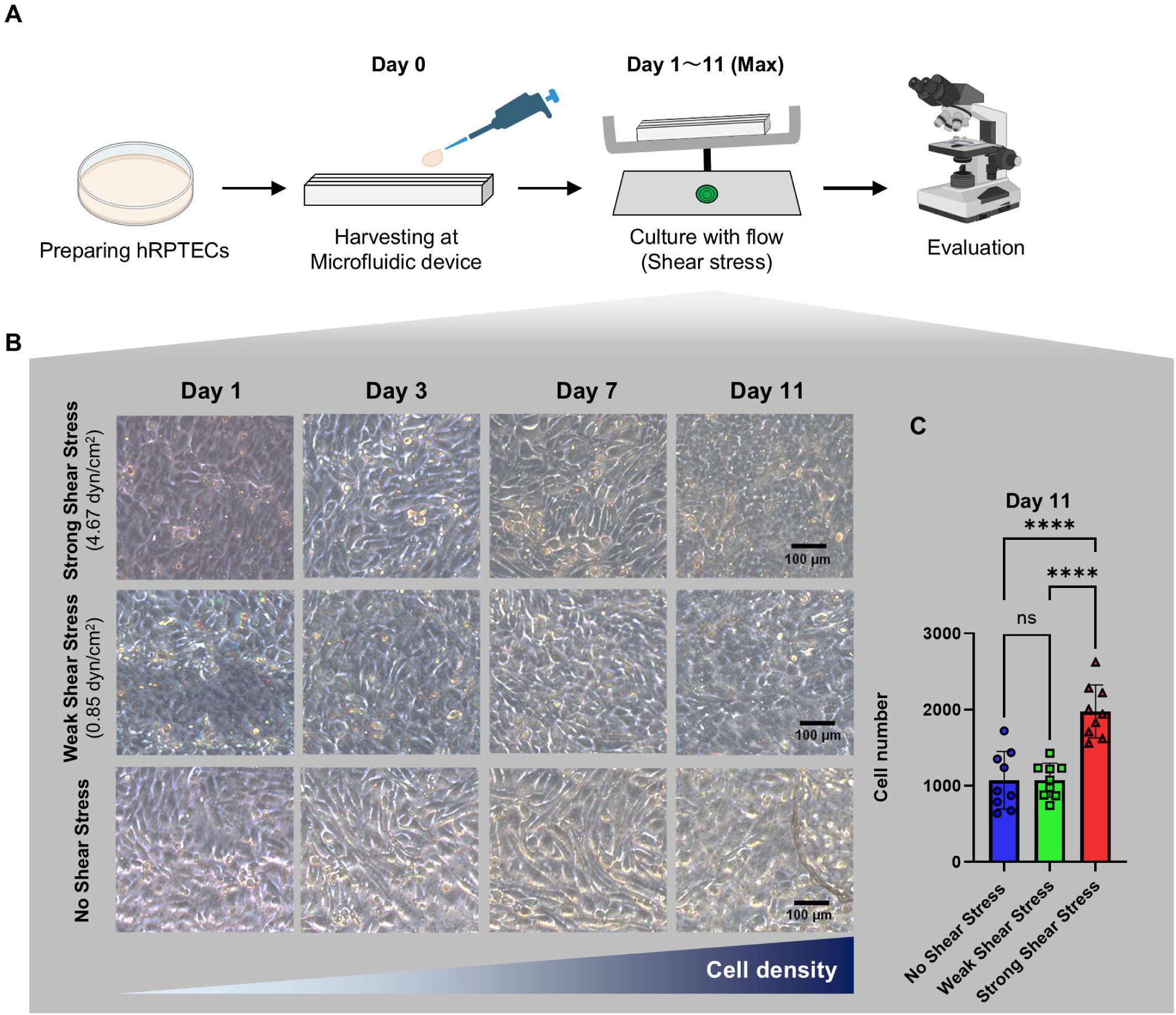
Physiological-level shear stress increases human renal proximal tubular epithelial cells (hRPTEC) density in a human proximal tubule microphysiological system (hPT-MPS). **(A)** Experimental workflow for hRPTEC seeding, rocker-driven shear stress culture, and microscopic evaluation. **(B)** Representative phase-contrast images of hRPTECs cultured under high, low, or no shear stress conditions at Days 1, 3, 7, and 11 (scale bars, 100 μm). **(C)** Quantification of cell number per field on Day 11 under static (no shear stress), low-shear stress, and high-shear stress conditions. Cells were manually counted from phase-contrast images acquired at 400× magnification (*n* = 9 per condition). Data are presented as mean ± standard error of the mean. *P* < 0.0001, static versus high shear; *p* < 0.0001, low shear versus high shear. Figure.2 was created in part using <u>BioRender.com</u>.

Phase-contrast imaging revealed a progressive increase in cell density under high-shear stress conditions from Day 1 to Day 11, whereas cells cultured under static (no shear stress) conditions exhibited comparatively lower confluency by Day 11 (**Figure 2B**). Low-shear stress conditions produced an intermediate phenotype, although the increases in cell density were less pronounced than those observed under high-shear stress conditions.

Quantitative analysis on Day 11 demonstrated that high-shear stress conditions significantly increased the number of cells per field compared with both static (no shear stress) and low-shear stress conditions (**Figure 2C**). No significant difference was observed between the static and low-shear stress conditions. These findings suggest that physiological-level rocker-driven shear stress enhances cell accumulation and supports epithelial tissue organization within the hPT-MPS.

### 3.3. Rocker-Driven Shear Stress Promotes Proximal Tubule-Associated Marker Expression and Epithelial Organization

To investigate whether shear stress promotes proximal tubule epithelial maturation, the expression of proximal tubule-associated markers and epithelial junction proteins was assessed.

Immunofluorescence analysis demonstrated that Na /K^+^-ATPase expression was increased under high-shear stress conditions (**Figure 3A**). Quantitative analysis confirmed significantly greater Na/K^+^-ATPase signal intensity on Day 11 under high-shear stress conditions compared with both Day 1 and static (no shear stress) controls (**Figure 3B**). Similarly, expression of the proximal tubule water channel AQP1 was markedly increased under high-shear stress conditions, with significantly greater signal intensity on Day 11 than under static conditions (**Figure 3C**).

**Figure 3.**
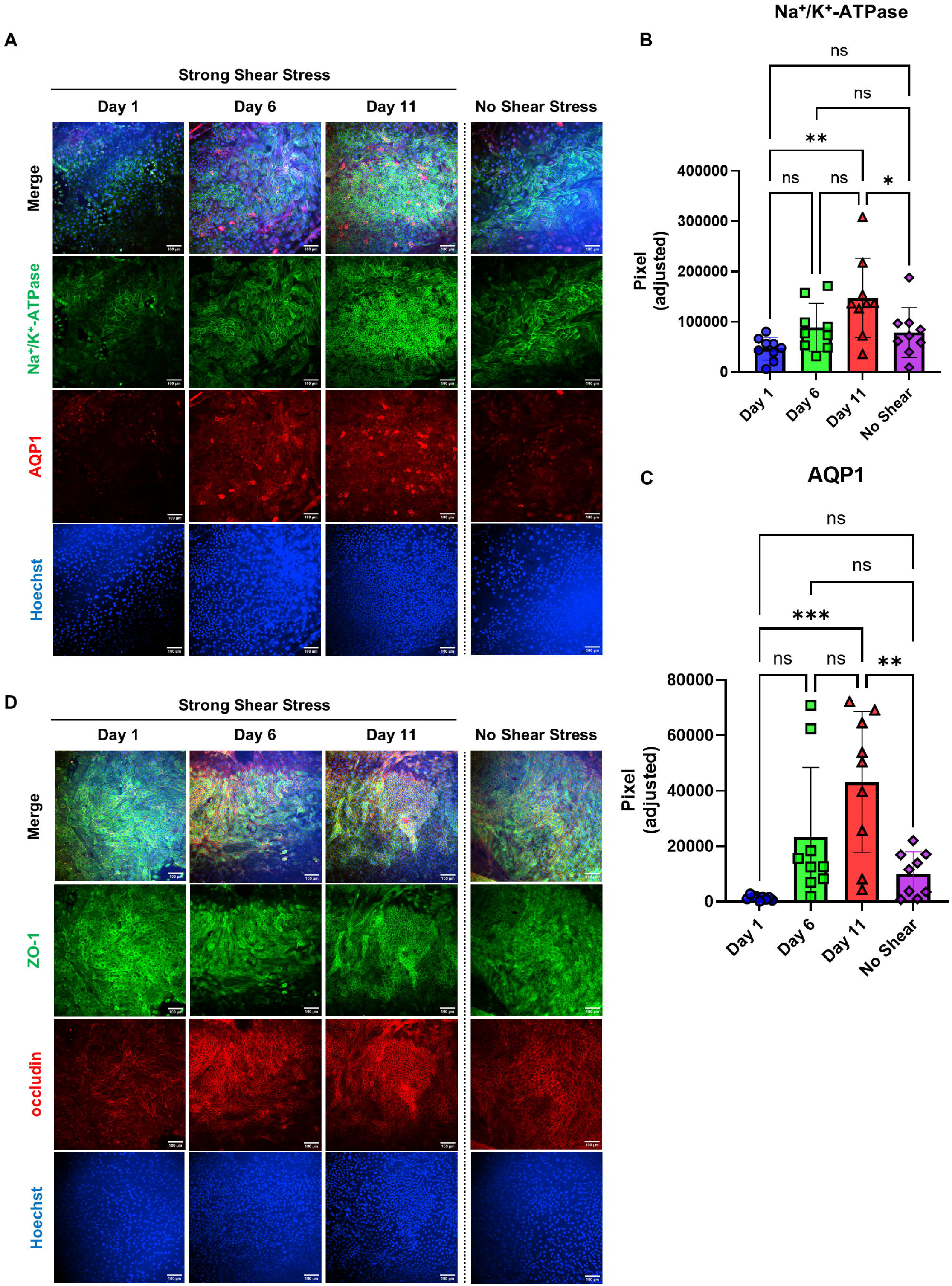
Rocker-driven shear stress promotes proximal tubule-associated marker expression and epithelial junctional organization. **(A)** Representative immunofluorescence images of Na^+^/K^+^-ATPase, aquaporin-1 (AQP1), and Hoechst in human renal proximal tubular epithelial cells (hRPTECs) cultured under high-shear stress conditions at Days 1, 6, and 11 and under static (no shear stress) conditions at Day 11 (scale bars, 100 μm). **(B)** Quantification of Na^+^/K^+^-ATPase fluorescence intensity (*n* = 9 from three independent experiments). Data are presented as mean ± standard error of the mean. *P* = 0.0019, Day 1 versus Day 11; *p* = 0.0492, Day 11 versus static. **(C)** Quantification of the aquaporin-1 (AQP1) fluorescence intensity (*n* = 9 from three independent experiments). Data are presented as mean ± standard error of the mean. *P* = 0.0002, Day 1 versus Day 11; p = 0.0031, Day 11 versus static. **(D)** Representative immunofluorescence images of zonula occludens-1 (ZO-1), occludin, and Hoechst in hRPTECs cultured under high-shear conditions at Days 1, 6, and 11 and under static (no shear stress) conditions at Day 11 (scale bars, 100 μm).

Next, epithelial junctional organization was assessed by immunostaining for ZO-1 and occludin. Under high-shear stress conditions, both ZO-1 and occludin staining exhibited more organized junctional localization than under static (no shear stress) conditions (**Figure 3D**). Collectively, these findings suggest that rocker-driven shear stress promotes the acquisition of maturation-associated features of hRPTECs, including enhanced expression of proximal tubule-associated markers and improved epithelial junctional organization.

### 3.4. RNA-seq Analysis Reveals Shear Stress-Associated Transcriptional Remodeling toward Epithelial Maturation

To investigate the molecular effects of shear stress, bulk RNA sequencing was performed on hRPTECs cultured under high-shear stressor static (no shear stress) conditions on Day 11 (**Figure 4A**). Genes encoding proteins assessed by immunofluorescence, including AQP1, ATP1A1, OCLN, and TJP1, showed a similar upward trend under shear stress, although these differences did not reach statistical significance in the RNA-seq analysis (**Supplementary Figure S2A**).

**Figure 4.**
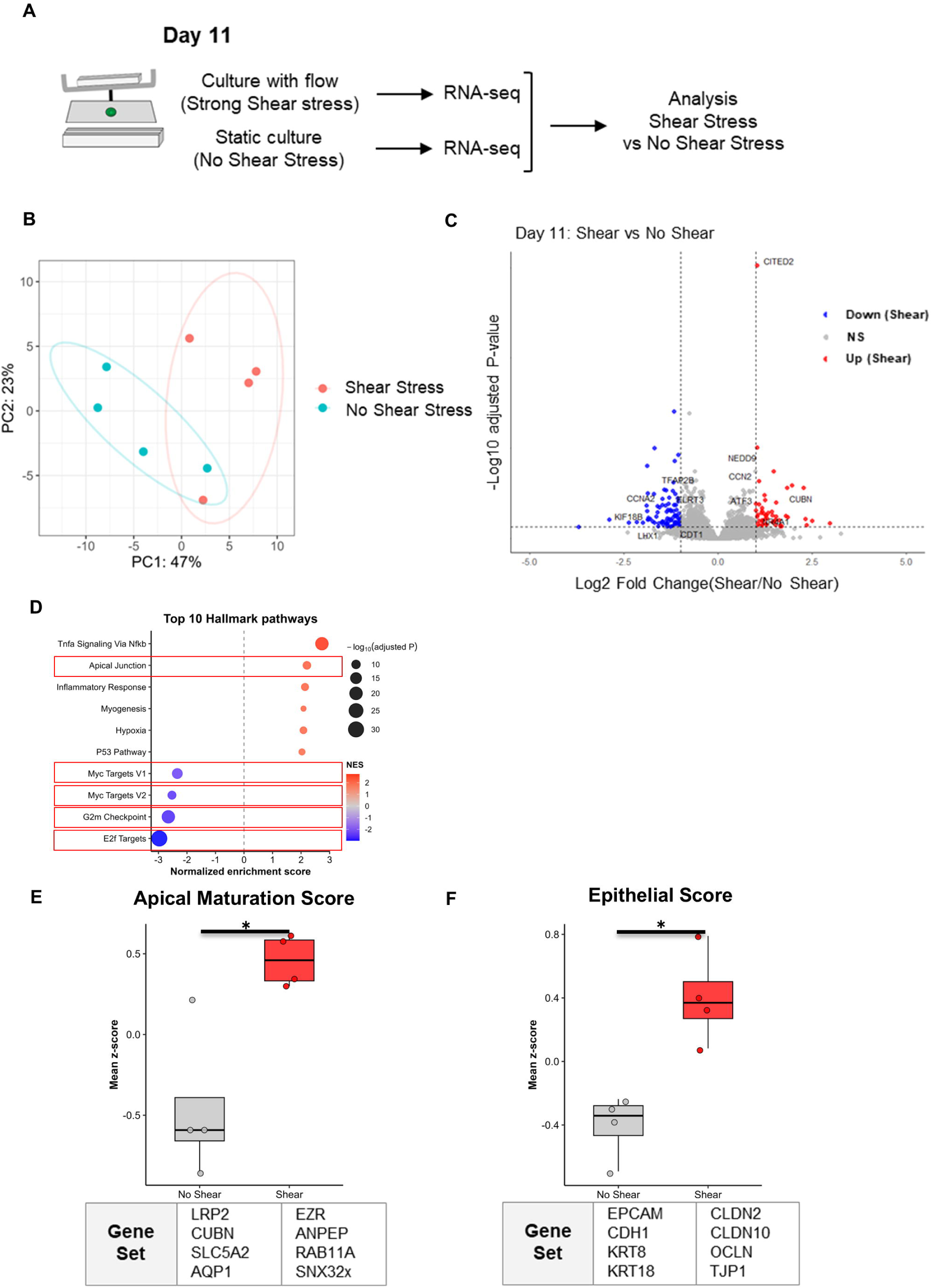
Shear stress induces transcriptional remodeling toward epithelial maturation in human renal proximal tubular epithelial cells (hRPTECs). **(A)** Experimental design for bulk RNA sequencing (RNA-seq) analysis of hRPTECs cultured under high or static (no shear stress) conditions on Day 11. **(B)** Principal component analysis of RNA-seq samples under high-shear stress and static (no shear stress) conditions. **(C)** Volcano plot showing differentially expressed genes between high-shear stress and static conditions on Day 11. **(D)** Hallmark gene set enrichment analysis of shear stress-associated transcriptional changes. **(E)** Apical maturation score calculated from the indicated proximal tubule-associated gene set under static and high-shear stress conditions (*n* = 4 per condition; *p* = 0.03). **(F)** Epithelial score calculated from the indicated epithelial marker gene set under static and high-shear stress conditions (*n* = 4 per condition; *p* = 0.03).

Principal component analysis revealed clear separation between high-shear and static (no shear stress) samples, indicating that shear stress induces distinct transcriptional profiles **(Figure 4B**). Differential gene expression analysis further identified shear stress-associated transcriptional changes (**Figure 4C**). Among genes upregulated under shear stress, *CUBN, CCN2, NEDD9,* and *CITED2* were detected. Notably, *CUBN*, which encodes the proximal tubule endocytic receptor cubilin, was significantly upregulated under shear stress conditions. *LRP2*, which encodes another proximal tubule-associated endocytic receptor, also showed an upward trend, although the difference did not reach statistical significance (**Supplementary Figure S2B**). Conversely, several cell cycle-associated genes were downregulated under shear stress, suggesting a shift away from a proliferative transcriptional state.

Gene set enrichment analysis revealed shear stress-associated enrichment of epithelial and apical surface-related pathways, along with alterations in proliferative and cell cycle-related pathways (**Figure 4D**). To further quantify epithelial maturation, we calculated composite scores based on predefined gene sets. The apical maturation score was significantly increased under shear stress when compared with static (no shear stress) conditions (*p* = 0.03; **Figure 4E**). Similarly, the epithelial score was significantly elevated under shear stress (*p* = 0.03; **Figure 4F**).

Together, these transcriptomic analyses support the concept that sustained rocker-driven shear stress promotes epithelial maturation-associated transcriptional remodeling in patient-derived hRPTECs.

### 3.5. Shear Stress Alters Cisplatin Responsiveness in the Human Patient-Derived Proximal Tubule Microphysiological System

To evaluate whether shear stress-conditioned hPT-MPS cultures recapitulate nephrotoxic injury responses, Day 11 cultures were exposed to increasing concentrations of cisplatin under shear or static (no shear stress) conditions for 48 hours (**Figure 5A**).

**Figure 5.**
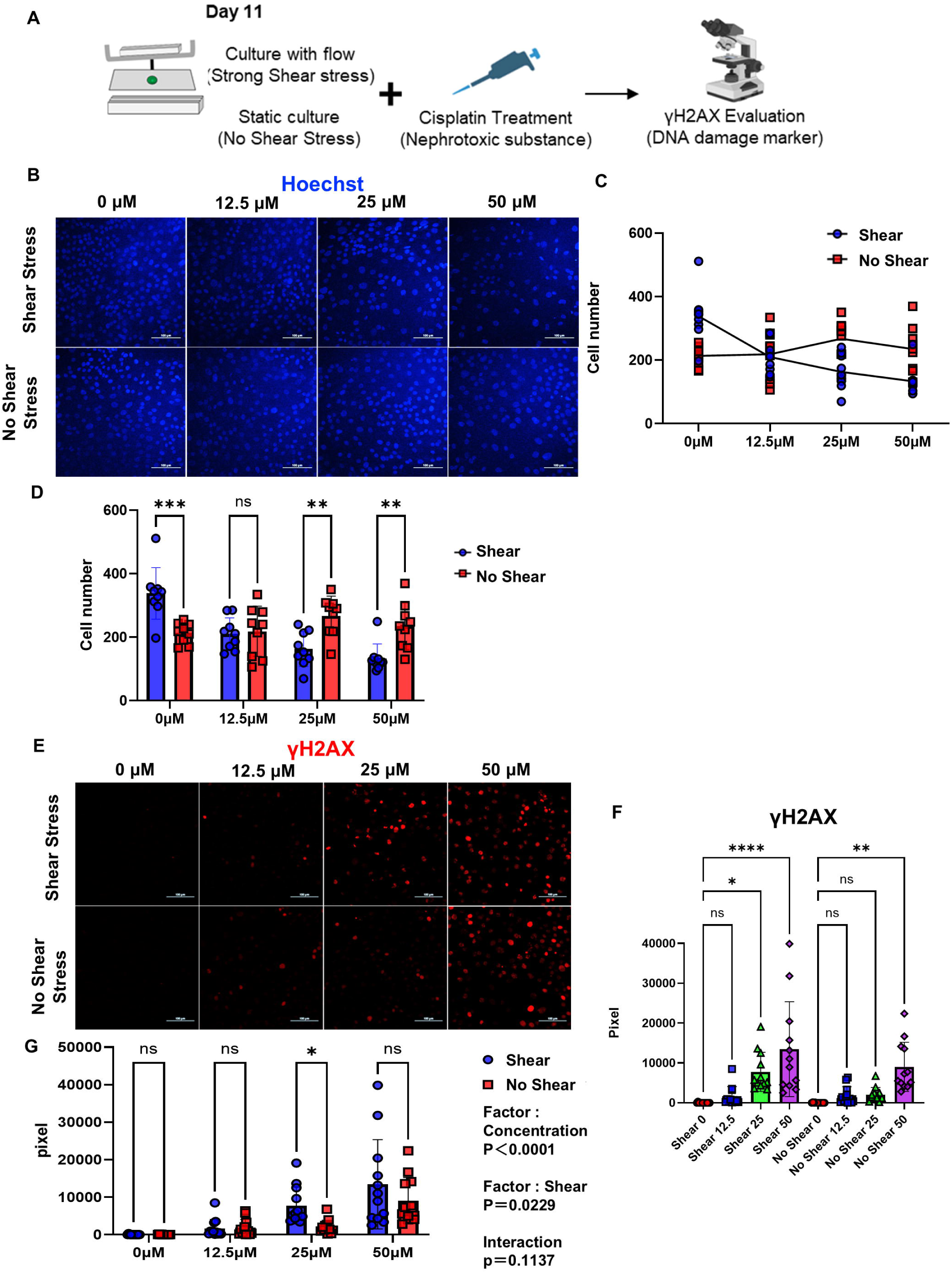
Shear stress alters cisplatin responsiveness in human proximal tubule microphysilogical system (hPT-MPS). **(A)** Experimental design for cisplatin treatment and γH2AX assessment in human renal proximal tubular epithelial cells (hRPTECs) cultured under high-shear stress or static (no shear stress) conditions. **(B)** Representative Hoechst images of hRPTECs treated with 0, 12.5, 25, or 50 μM cisplatin under shear stress or static conditions (scale bars, 100 μm). **(C)** Cell number per field after cisplatin treatment under high-shear stress or static conditions, shown as a concentration-response plot. **(D)** Quantification of residual cell numbers after cisplatin treatment. The same dataset as in Figure 5C is replotted as grouped bars to directly compare high-shear stress and static conditions at each cisplatin concentration. Data are presented as mean ± standard error of the mean (*n* = 9 from three independent experiments). *P* = 0.0003, 0 μM high-shear stress versus static; *p* = 0.0027, 25 μM high-shear versus static; *p* = 0.0038, 50 μM high-shear stress versus static. **(E)** Representative γH2AX immunofluorescence images of hRPTECs treated with 0, 12.5, 25, or 50 μM cisplatin under high-shear stress or static conditions (scale bars, 100 μm). **(F)** Quantification of γH2AX signal intensity across cisplatin concentrations under high-shear and static conditions. Data are presented as mean ± standard error of the mean (*n* = 12 from four independent experiments). Under high-shear stress conditions, *p* = 0.0104 for 25 μM versus 0 μM and *p* < 0.0001 for 50 μM versus 0 μM. Under static conditions, *p* = 0.0014 for 50 μM versus 0 μM. **(G)** Comparison of γH2AX signal intensity between high-shear stress and static conditions at each cisplatin concentration. The same dataset as in Figure 5F is replotted to directly compare conditions. Data are presented as mean ± standard error of the mean. *P* = 0.0343, 25 μM high-shear stress versus static. Two-way analysis of variance showed significant main effects of cisplatin concentration and shear condition, with *p* < 0.0001 and *p* = 0.0229, respectively, with no significant interaction (*p* = 0.1137). Figure.5 was created in part using <u>BioRender.com</u>.

**Figure 6.**
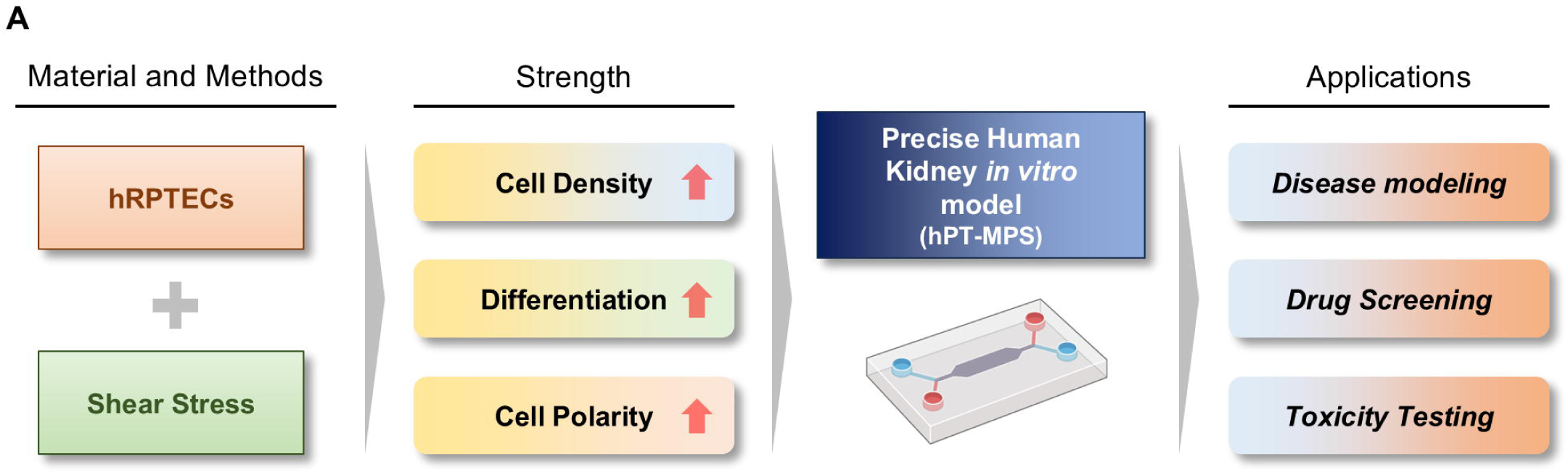
The patient-derived human proximal tubule microphysiological system (hPT-MPS) provides a pump-free platform for modeling flow-dependent proximal tubule phenotypes. Schematic summary of the human proximal tubule microphysiological system (hPT-MPS), consisting of human renal proximal tubular epithelial cells (hRPTECs), a rocker-driven shear stress platform, and a microfluidic device. The platform supports potential applications in disease modeling, drug screening, and toxicity testing. Figure 6 was created in part using BioRender.com.

First, the nuclei were stained with Hoechst, and the residual cell number was quantified after cisplatin treatment (**Figure 5B**). The same dataset was presented in two complementary formats: a concentration-response plot illustrating the overall trend across cisplatin doses (**Figure 5C**) and a grouped bar plot directly comparing shear and static (no shear stress) conditions at each concentration (**Figure 5D**). In the absence of cisplatin, cell numbers were higher under shear conditions than under static conditions. This difference was no longer apparent at 12.5 μM cisplatin and was reversed at 25 μM and 50 μM, with fewer cells remaining under shear conditions than under static conditions **(Figures 5C and 5D**). Collectively, these findings suggest that shear-conditioned epithelia exhibit increased susceptibility to cisplatin-induced cell loss.

Next, DNA damage was assessed by immunostaining for γH2AX, a marker of DNA double-strand breaks [15]. Cisplatin induced a concentration-dependent increase in the γH2AX signal under both culture conditions (**Figures 5E and 5F**). The same γH2AX quantification dataset was then replotted to directly compare shear stress and static (no shear stress) conditions at each cisplatin concentration (**Figure 5G**). Under shear conditions, the γH2AX signal was significantly increased at 25 μM and 50 μM cisplatin compared with untreated controls. Under static conditions, a significant increase was observed only at 50 μM cisplatin. Direct comparison between shear stress and static (no shear stress) conditions showed a significantly higher γH2AX signal under shear conditions at 25 μM cisplatin, whereas no significant differences were observed at 0, 12.5, or 50 μM (**Figure 5G**). Two-way analysis of variance revealed significant main effects of cisplatin concentration and shear condition, without a significant interaction between two factors. Collectively, these findings indicate that shear stress alters cisplatin responsiveness in the hPT-MPS and support its utility as a platform for evaluating drug-induced tubular injury.

## 4. Discussion

In this study, a pump-free, patient-derived hPT-MPS was developed by integrating hRPTECs isolated from surgically resected kidney tissue with a porous membrane-based microfluidic device and a rocker-driven shear stress platform. Using this system, physiological-level shear stress increased epithelial density, enhanced expression of proximal tubule-associated markers, improved epithelial junctional organization, and induced transcriptomic changes consistent with epithelial maturation. Shear-conditioned epithelia also exhibited altered responsiveness to cisplatin-induced injury, as reflected by reduced residual cell number and increased γH2AX signal. Collectively, these findings suggest that mechanical conditioning is a key determinant of proximal tubule epithelial phenotype and injury response in patient-derived human kidney cells.

The role of shear stress in proximal tubule biology is well supported by previous studies demonstrating that tubular epithelial cells are mechanosensitive. *In vivo*, proximal tubule cells are continuously exposed to luminal flow and fluid shear stress, which are sensed through apical structures such as primary cilia, microvilli, the glycocalyx, and the actin cytoskeleton [16]. These mechanosensory pathways regulate epithelial architecture, cytoskeletal organization, endocytic capacity, and transport-related functions [16, 17]. Consistent with this framework, we observed increased Na^+^/K^+^-ATPase and AQP1 expression, more organized ZO-1 and occludin localization, and higher apical maturation and epithelial scores under shear stress. Although direct transporter activity was not assessed in this study, these structural and transcriptional changes support the concept that shear stress promotes maturation-associated epithelial features in patient-derived hRPTECs.

Our findings are also consistent with previous reports highlighting the importance of fluid shear stress in proximal tubule models. Birdsall and Hammond summarized evidence that shear stress promotes structural remodeling, microvilli formation, albumin handling, and drug transporter expression in proximal tubular cell models, while also noting that shear stress conditions vary substantially across platforms and should be carefully reported to ensure reproducibility [18]. In this context, our hPT-MPS offers a simple and scalable approach for applying shear stress without the need for individual pumps or tubing. The rocker-driven configuration reduces technical complexity and enables parallel operation of multiple devices, which is advantageous for nephrotoxicity testing and platform-based screening.

A notable finding of this study was that shear-conditioned epithelia exhibited altered responsiveness to cisplatin. Cisplatin treatment reduced residual cell number more prominently under shear stress, and γH2AX signal was significantly higher under shear stress conditions at 25 μM cisplatin. These results suggest that the epithelial state induced by shear stress influences the apparent susceptibility of proximal tubular cells to nephrotoxic injury. Cisplatin nephrotoxicity involves proximal tubular accumulation, DNA damage, mitochondrial dysfunction, oxidative stress, and inflammatory signaling, with transporters such as organic cation transporter 2 (OCT2) and multidrug and toxin extrusion protein 1 contributing to renal cisplatin handling [19, 20]. In our dataset, OCT2 and multidrug and toxin extrusion protein 1 did not show marked transcriptional increases (**Supplementary Figure S3A**), and OCT2 immunostaining did not reveal substantial differences between conditions (**Supplementary Figure S3B**). In contrast, copper transporter 1, which has been implicated in cisplatin uptake [21], was increased under shear conditions (**Supplementary Figure S3C**). Although this observation does not establish causality, it raises the possibility that alternative uptake pathways or maturation-associated changes in the epithelial state may contribute to the enhanced cisplatin response.

Several limitations of this study should be acknowledged. First, although immunostaining and transcriptomic analyses supported maturation-associated changes under shear stress, we did not perform direct functional assays of transporter activity, albumin uptake, solute reabsorption, or barrier permeability. Second, intracellular cisplatin accumulation was not directly measured; therefore, it remains unclear whether the enhanced injury response under shear stress reflects differences in drug uptake, intracellular handling, or downstream DNA damage signaling. Third, shear stress was estimated theoretically, and the dynamic time-varying nature of rocker-driven bidirectional flow was not experimentally quantified. Fourth, because the platform is based on primary patient-derived cells, donor-to-donor variability may influence baseline epithelial phenotype and toxicological responses. While this variability is an inherent feature for human-relevant models, it will require validation in larger donor cohorts.

Despite these limitations, this study establishes a practical patient-derived hPT-MPS that can be operated without individual pumps and used to investigate flow-dependent proximal tubule phenotypes. Future studies incorporating functional transport assays, direct measurements of drug accumulation, and additional nephrotoxic compounds will further strengthen the utility of this platform. In addition, the bilayer structure of the device enables future incorporation of stromal, endothelial, or immune cell types in the lower compartment, allowing more complex modeling of the renal microenvironment. The patient-derived nature of this system may also support the development of disease-specific MPS models using cells obtained from patients with chronic kidney disease or other kidney disorders.

## Data Availability

All data produced in the present study are available upon reasonable request to the authors

## Funding

This work was supported by JST SPRING (JMPJSP2180), Japan Science and Technology Agency (to Yuta Sekiguchi); the Leading Initiative for Excellent Young Researchers, Ministry of Education, Culture, Sports, Science and Technology (to Yutaro Mori); JSPS KAKENHI Grant-in-Aid for Research Activity Start-up (22K20881 to Yutaro Mori), Grant-in-Aid for Scientific Research (B) (24K03249 to Yutaro Mori), Grant-in-Aid for Transformative Research Areas (25H01351 to Hirokazu Kaji), Fund for the Promotion of Joint International Research (24KK0203 to Hirokazu Kaji), and Grant-in-Aid for Scientific Research (B) (to Yuji Nashimoto); the Innovation Idea Contest, Tokyo Medical and Dental University (TMDU) (2022 to Yutaro Mori and 2023 to Yuki Nakao); the Next Generation Researcher Training Unit, TMDU (to Yutaro Mori); the Priority Research Areas Grant, TMDU (to Yutaro Mori); the Uehara Memorial Foundation (to Yutaro Mori); the MSD Life Science Foundation Research Grant for Lifestyle-related Diseases (to Yutaro Mori); the Takeda Science Foundation Medical Research Grant (to Yutaro Mori); Academic Support from Bayer Yakuhin, Ltd. (to Yutaro Mori); the Kato Memorial Bioscience Foundation (to Yutaro Mori); The Waksman Foundation of Japan Inc. (to Yutaro Mori); the Pharmacodynamics Research Society (to Yutaro Mori); the Japan Agency for Medical Research and Development (AMED) (JP25ek0310028h0001 to Yutaro Mori); and Fusion Oriented Research for disruptive Science and Technology (FOREST), Japan Science and Technology Agency (JPMJFR245N to Yutaro Mori and JPMJFR234S to Yuji Nashimoto).

## Acknowledgments

We are deeply grateful to the study participants who generously consented to donate pieces of their resected kidneys for this research.

## Data Availability Statement

Information and requests for resources and reagents should be directed to and will be fulfilled by the Lead Contact Yutaro Mori.

## Author Contributions

Yuta Sekiguchi, Ayumi Suzuki, Yuki Nakao, Yuji Nashimoto, Takeshi Hori, Makiko Mori, and Afreen Fatima performed experiments, collected and analyzed data, and wrote the manuscript. Yuta Sekiguchi, Yuki Nakao, Makiko Mori, Ryota Shindoh, Iori Morita, and Yutaro Mori established hRPTECs. Shintaro Mandai, Tamami Fujiki, Hiroaki Kikuchi, Yohei Arai, Fumiaki Ando, Koichiro Susa, Takayasu Mori, and Eisei Sohara supported the data analysis. Yuta Sekiguchi performed the analysis of the bulk RNA-seq data. Ayumi Suzuki, Yuji Nashimoto, and Hirokazu Kaji prepared the microfluidic device and provided technical support. Yuma Waseda, Soichiro Yoshida, and Yasuhisa Fujii resected the patients’ kidneys as standard treatment for malignant diseases. Yutaro Mori, Yuji Nashimoto, and Hirokazu Kaji developed the experimental strategy, supervised the project, and edited the manuscript. All authors discussed the results and implications and commented on the manuscript.

## Conflict of Interest

The authors declare that they have no conflicts of interest.

---

**Supplementary Figure S1.**
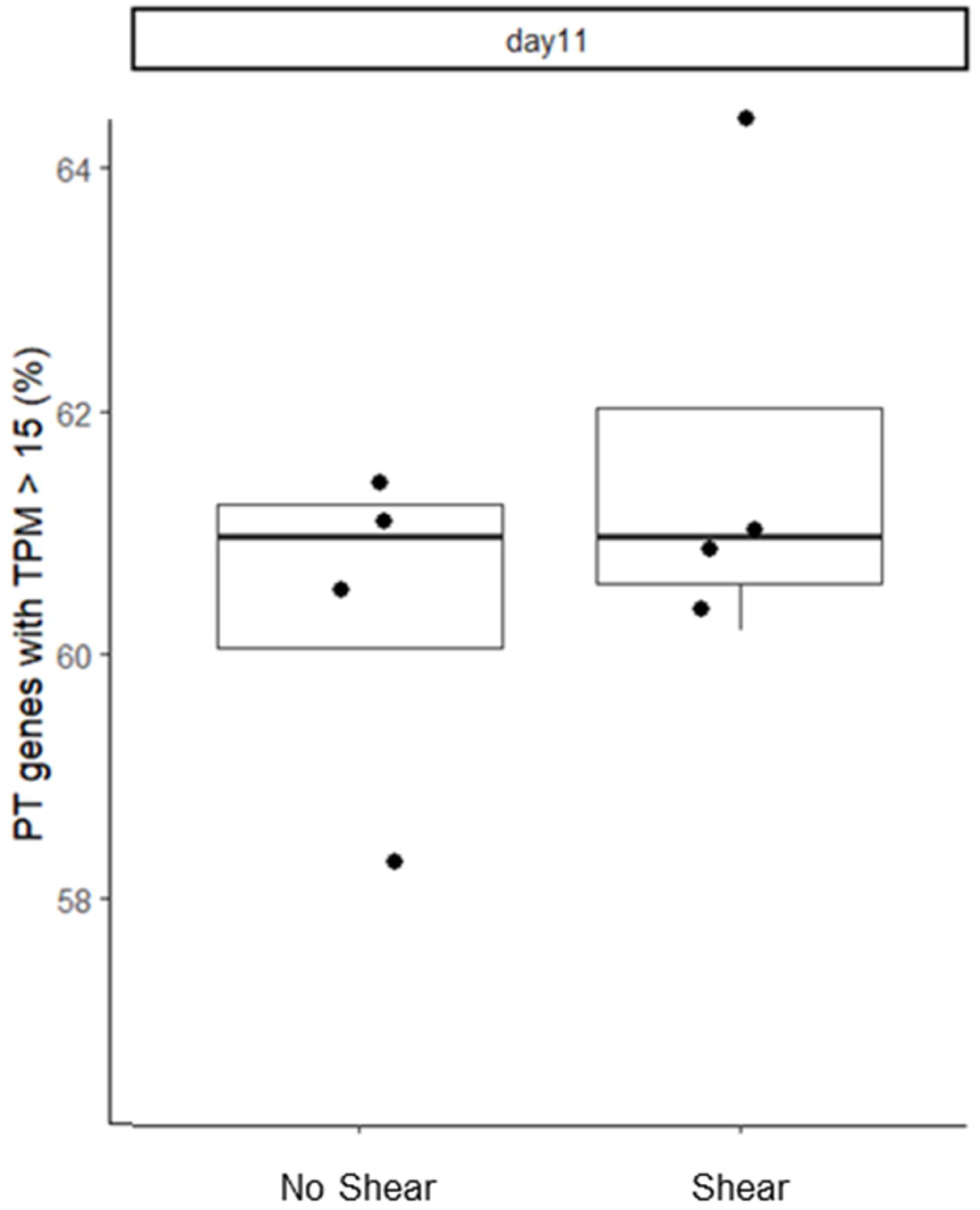
hRPTECs retain broad proximal tubule-associated transcriptional features in the hPT-MPS. Percentage of previously reported proximal tubule-associated genes expressed at TPM > 15 in hRPTECs cultured under no shear stress or shear stress conditions at Day 11.

**Supplementary Figure S2.**
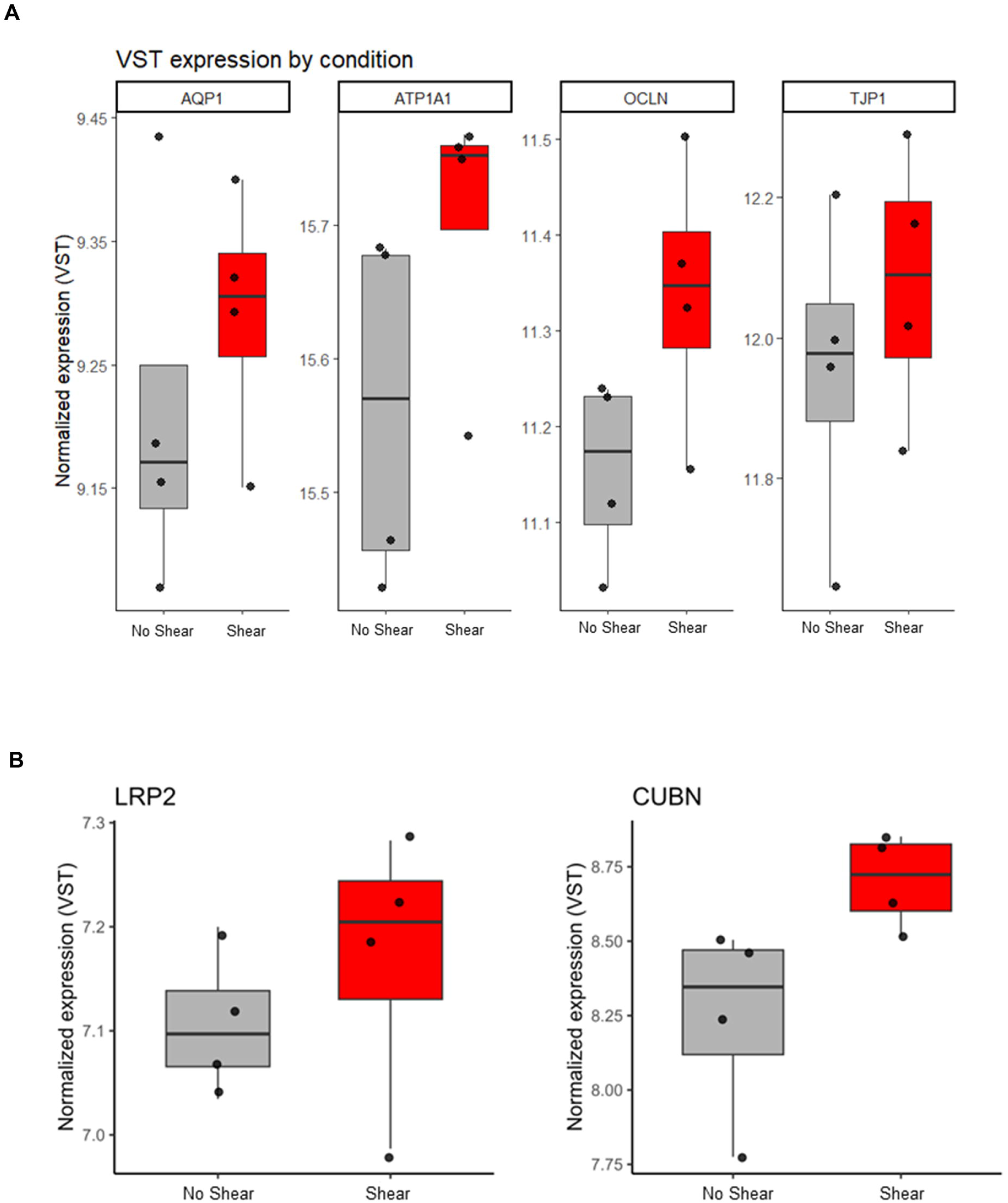
Shear stress increases selected proximal tubule and epithelial marker transcripts. (A) Variance-stabilized expression levels of AQP1, ATP1A1, OCLN, and TJP1 under no shear stress and shear stress conditions. (B) Variance-stabilized expression levels of LRP2 and CUBN under no shear stress and shear stress conditions.

**Supplementary Figure S3.**
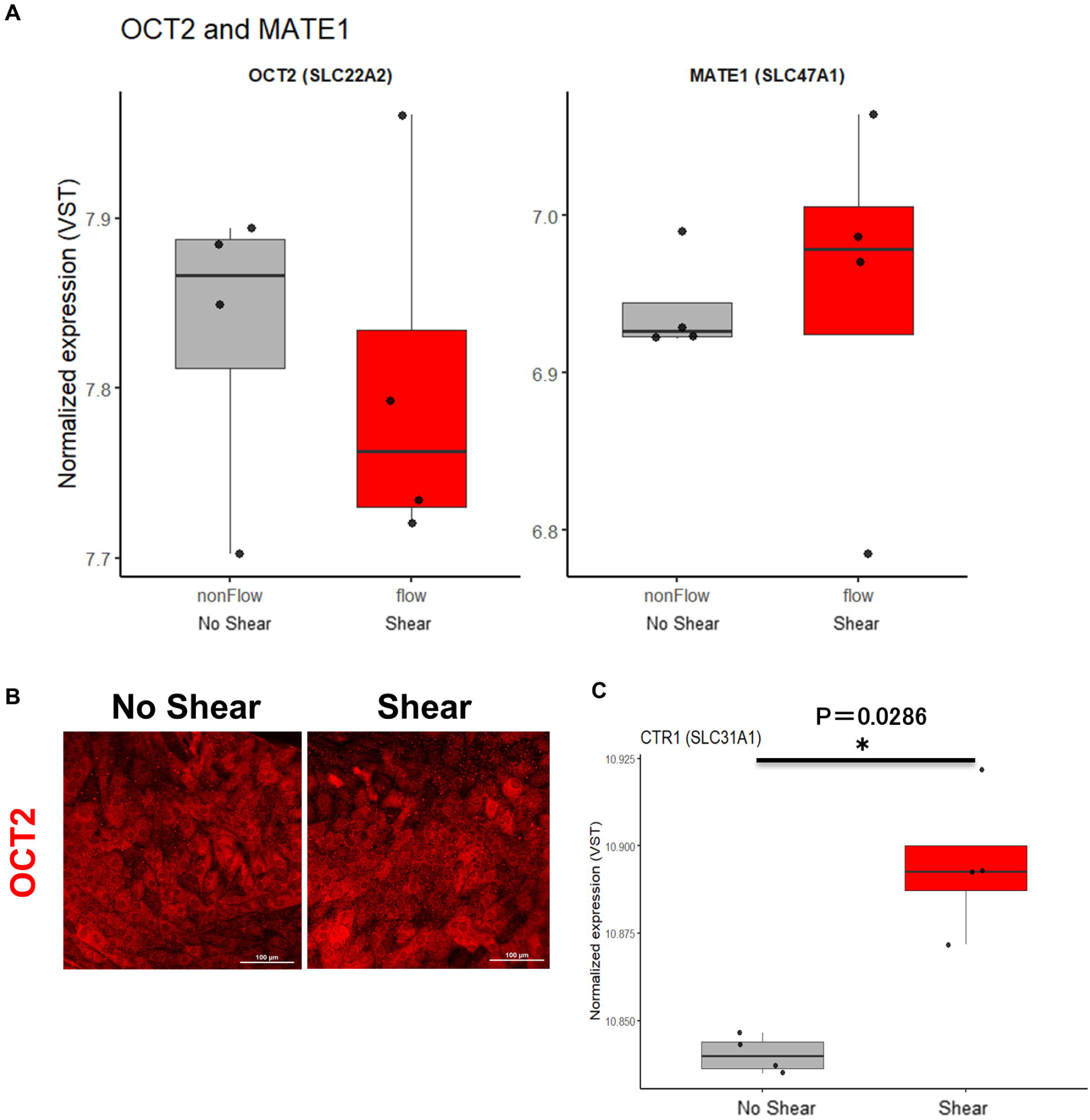
Shear stress alters the expression of cisplatin-associated transporters. (A) RNA-seq expression levels of OCT2/SLC22A2 and MATE1/SLC47A1 under no shear stress and shear stress conditions. (B) Representative immunofluorescence images of OCT2 under no shear stress and shear stress conditions. Scale bars, 100 µm. (C) RNA-seq expression of CTR1/SLC31A1 under no shear stress and shear stress conditions. p = 0.0286.

## Notes

### Competing Interest Statement

The authors have declared no competing interest.

### Author Declarations

The Institutional Review Board of the Institute of Science Tokyo Hospital gave ethical approval for this work (approval no. M2022-005)

## References

1. Poh, W.T. and J. Stanslas, The new paradigm in animal testing - “3Rs alternatives”. Regul Toxicol Pharmacol, 2024. 153: p. 105705.

2. Carratt, S.A., et al., An industry perspective on the FDA Modernization Act 2.0/3.0: potential next steps for sponsors to reduce animal use in drug development. Toxicol Sci, 2025. 203(1): p. 28–34.

3. Pound, P. and M. Ritskes-Hoitinga, Is it possible to overcome issues of external validity in preclinical animal research? Why most animal models are bound to fail. J Transl Med, 2018. 16(1): p. 304.

4. Loewa, A., J.J. Feng, and S. Hedtrich, Human disease models in drug development. Nat Rev Bioeng, 2023: p. 1–15.

5. Leenaars, C.H.C., et al., Animal to human translation: a systematic scoping review of reported concordance rates. J Transl Med, 2019. 17(1): p. 223.

6. Van Norman, G.A., Limitations of Animal Studies for Predicting Toxicity in Clinical Trials: Is it Time to Rethink Our Current Approach? JACC Basic Transl Sci, 2019. 4(7): p. 845–854.

7. Marshall, L.J., et al., Poor Translatability of Biomedical Research Using Animals - A Narrative Review. Altern Lab Anim, 2023. 51(2): p. 102–135.

8. Ineichen, B.V., et al., Analysis of animal-to-human translation shows that only 5% of animal-tested therapeutic interventions obtain regulatory approval for human applications. PLoS Biol, 2024. 22(6): p. e3002667.

9. Loghman-Adham, M., et al., Detection and management of nephrotoxicity during drug development. Expert Opin Drug Saf, 2012. 11(4): p. 581–96.

10. Homan, K.A., et al., Flow-enhanced vascularization and maturation of kidney organoids in vitro. Nat Methods, 2019. 16(3): p. 255–262.

11. Banan Sadeghian, R., et al., Cells sorted off hiPSC-derived kidney organoids coupled with immortalized cells reliably model the proximal tubule. Commun Biol, 2023. 6(1): p. 483.

12. Ross, E.J., et al., Three dimensional modeling of biologically relevant fluid shear stress in human renal tubule cells mimics in vivo transcriptional profiles. Sci Rep, 2021. 11(1): p. 14053.

13. Ichimura, T., et al., Kidney injury molecule-1 is a phosphatidylserine receptor that confers a phagocytic phenotype on epithelial cells. J Clin Invest, 2008. 118(5): p. 1657–68.

14. Khundmiri, S.J., et al., Transcriptomes of Major Proximal Tubule Cell Culture Models. J Am Soc Nephrol, 2021. 32(1): p. 86–97.

15. Mah, L.J., A. El-Osta, and T.C. Karagiannis, gammaH2AX: a sensitive molecular marker of DNA damage and repair. Leukemia, 2010. 24(4): p. 679–86.

16. Weinbaum, S., et al., Mechanotransduction in the renal tubule. Am J Physiol Renal Physiol, 2010. 299(6): p. F1220–36.

17. Raghavan, V. and O.A. Weisz, Flow stimulated endocytosis in the proximal tubule. Curr Opin Nephrol Hypertens, 2015. 24(4): p. 359–65.

18. Birdsall, H.H. and T.G. Hammond, Role of Shear Stress on Renal Proximal Tubular Cells for Nephrotoxicity Assays. J Toxicol, 2021. 2021: p. 6643324.

19. McSweeney, K.R., et al., Mechanisms of Cisplatin-Induced Acute Kidney Injury: Pathological Mechanisms, Pharmacological Interventions, and Genetic Mitigations. Cancers (Basel), 2021. 13(7).

20. Sprowl, J.A., et al., Cisplatin-induced renal injury is independently mediated by OCT2 and p53. Clin Cancer Res, 2014. 20(15): p. 4026–35.

21. Pabla, N., et al., The copper transporter Ctr1 contributes to cisplatin uptake by renal tubular cells during cisplatin nephrotoxicity. Am J Physiol Renal Physiol, 2009. 296(3): p. F505–11.

