## Supplementary method for "Pump-Free Patient-Derived Human Proximal Tubule Microphysiological System for Modeling Flow-Dependent Epithelial Maturation and Cisplatin Injury"

**Supplementary Methods (The details of RNA-seq analysis)**

**RNA extraction, library preparation, and sequencing**

Total RNA was extracted in Sepasol-RNA I Super G (Nacalai Tesque, Japan), followed by purification using the Direct-zol RNA Miniprep Plus Kit (Zymo Research, CA, USA) according to the manufacturer’s protocol. RNA integrity was assessed using an Agilent Bioanalyzer by Rhelixa Inc, and all samples had RIN values greater than 7.5. RNA-seq library preparation and sequencing were also outsourced to Rhelixa Inc. using NEBNext kits, libraries were prepared in a non-strand-specific manner. Sequencing was performed on an Illumina NovaSeq X Plus system, generating 2 × 150 bp paired end reads (PE150), with approximately 6 giga bases of data and 40 million reads (20 million read pairs) per sample.

**Read preprocessing and quality control**

Raw reads were trimmed and filtered using fastp (v0.22.0) with the options: -q 20 -u 30 -n 10 -l 30 -w 8. Quality control was performed before and after trimming using FastQC (v0.12.1), and consolidated quality reports were generated using MultiQC (v1.28).

**Transcript quantification and annotation**

Transcript-level quantification was performed using Salmon (v1.10.3). The transcriptome index was built using GENCODE v45 (GRCh38). Gene-level count matrices were generated using the tximport package with the option countsFromAbundance = "lengthScaledTPM" to incorporate transcript length normalization.
